# Design, Implementation, and Evaluation of a Shadowing Program for Medical Students in the Basic Sciences Phase

**DOI:** 10.64898/2026.06.10.26355363

**Authors:** Athar Omid, Tahereh Changiz, Samane Ghasemi, Zahra Khodadoustan, Kian Heshmat, Aref Arefan, Mohammad Hossein Fazel Harandi, Mohammad Yousefi

## Abstract

**Introduction:** Shadowing, as an educational method based on active observation, can foster a realistic understanding of professional roles and enhance the communication skills of medical students. This study aimed to design, implement, and evaluate a shadowing program for basic sciences medical students.

**Methods:** This development study was conducted based on the ADDIE model in five phases. The study population consisted of 799 medical students in semesters 2 to 5. The stages included Analysis (determining needs through literature review and expert panels), Design (specifying learning environments and evaluation methods), Development (preparing guides and educational tools), Implementation (within the Medical Ethics course), and Evaluation (using questionnaires and reflection forms).

**Findings:** This study aimed to design and evaluate an educational shadowing program based on the ADDIE model. In the Analysis phase, the profiles of 799 students and learning objectives were determined. In the Design phase, a structured program for four types of shadowing was designed. In the Development phase, all guides and educational tools were prepared. In the Implementation phase, the program was carried out with complete coverage and adherence to ethical considerations. Finally, the program evaluation showed that “Motivation to become a good physician” (3.75-3.95) and “Enhancing empathy” (3.50-3.94) received the highest scores, while “Increasing understanding of the basic science-clinical connection**”** (2.53-2.89) and “Willingness to attend on holidays” (1.87-2.31) received the lowest scores.

**Conclusion:** The findings indicate that implementing the shadowing program is an effective method for strengthening the professional attitudes and academic motivation of medical students. However, the program did not significantly improve students’ perception of the basic science-clinical connection, indicating a need for curricular refinement. The continuation and extension of this program to other levels and fields of medical sciences are recommended.

## Introduction

One of the persistent challenges in undergraduate medical education is bridging the gap between preclinical coursework and clinical practice. Students in the basic sciences phase often struggle to visualize how foundational concepts translate into patient care, leading to reduced motivation and uncertainty (1). However, most studies have merely described this gap rather than critically examining why preclinical curricula fail to integrate clinical relevance (2, 3). Consequently, early exposure to clinical environments has become increasingly important.

Among educational approaches facilitating early exposure, observational learning has gained attention. Rooted in Bandura’s social learning theory (4), recent evidence questions whether passive observation without structured reflection produces measurable behavioral change (5, 6). Liberati (6) showed that observation alone leads to superficial understanding unless combined with pre-briefing and debriefing—a finding largely ignored in shadowing program designs.

One observational method is “shadowing.” Many curricula have adopted shadowing as a low-stakes strategy for introducing students to clinical settings (4). However, a critical review reveals that most published shadowing studies rely exclusively on post-intervention perception surveys, with no objective learning measures or control groups (7 ,5). Kitsis & Goldsammler (7) called for rigorous controlled designs over a decade ago, yet this gap persists (5, 8). Furthermore, comparative studies examining which shadowing type yields the greatest benefit are virtually absent (10 ,9).

In the basic sciences stage, shadowing may help students understand the medical profession and prevent future burnout (11). Nevertheless, the evidence base for preclinical shadowing is remarkably thin. Of 19 studies cited in a recent systematic review, only two focused on preclinical students, neither employing pre-post or controlled designs. A key unanswered question is whether basic sciences students—lacking clinical knowledge—can meaningfully benefit from shadowing or may instead experience confusion (5). Moreover, Dyrbye & Shanafelt’s (11) burnout study involved practicing physicians, not medical students, with no causal link tested for preclinical shadowing. Given the importance of this subject and the necessity of familiarizing basic sciences medical students with their field and professional future, this study aimed to design, implement, and evaluate a shadowing program for these students at Isfahan University of Medical Sciences.

## Methods

This was a development study conducted in five phases Analysis, Design, Development, Implementation, and Evaluation according to the stages of the ADDIE instructional design model (12). However, to address methodological limitations common in shadowing research (7 ,5) , we explicitly acknowledge the absence of a control group and a pre-post design, and we supplement descriptive statistics with inferential analyses where feasible.

### Research Population and Setting

The target population of this research consisted of all basic science’s medical students in semesters 2 to 5 (n=799) during the second semester of the 2023-2024 academic year. Twelve teaching hospitals of the university and comprehensive health centers throughout Isfahan province were selected as the program implementation settings.

The study implementation process, based on the phases of the ADDIE instructional design model, was as follows:

#### 1. Analysis

In this phase, audience analysis was performed. The gender, age, and number of learners were determined using the “Ham ava” educational system. Since this program was offered to students across four semesters in the basic sciences level, data for medical students in semesters 2 to 5 for the February 2024 semester were extracted. Additionally, based on the educational curriculum, the courses and training completed by this student group were reviewed. As the program was integrated into the four Medical Ethics I, II, III, and IV courses across four semesters, all relevant course information for the students was extracted.

To determine the needs and learning objectives, a literature review was first conducted. Based on this review, potential objectives for the shadowing program were identified. The literature review was performed in PubMed, ISI Web of Science, and Scopus databases. The search utilized a combination of the keywords “shadowing,” “patient shadowing,” “nurse shadowing,” “physician/doctor shadowing” with “medical education,” “health education,” and “education.”

The results of this literature review were presented in a focus group attended by the Vice Dean of General Medical Education of the faculty, two medical education specialists, two hospital education deputies, and two intern/extern students. After a general explanation of the shadowing program and subsequent discussion, the learning objectives in this program for each semester were determined. This meeting also defined *who* would be shadowed in each semester. Considering that the students in the first semester had no prior hospital experience, nurse shadowing was chosen for them so that they could have a more beneficial shadowing experience under the supervision of the nursing team. In semester 2, the experience of being at the patient’s bedside was selected; in semester 3, patient shadowing; in semester 4, intern shadowing; and in semester 5, health care provider shadowing.

#### 2. Program design

In this phase, the main framework and structure of the shadowing program were designed in detail during expert panel meetings attended by the Vice Dean of General Medical Education of the faculty, two medical education specialists, two hospital education deputies, and two intern/extern students. The design ensured that for each academic semester, considering the specific readiness level and educational objectives of that semester, the learning environment, educational approach, student assessment method, and program evaluation method were separately and appropriately defined. The final output of this design process was the creation of a precise operational plan for implementation.

#### 3. Development

In this phase, the educational content and required tools for the program were produced. The following actions were taken

- **Development of guides:** Educational guides were prepared for teaching assistants, those being shadowed (nurses, interns, on-call physicians, health care providers), supervisors, hospital education deputies, and the students themselves. These guides were in the form of a poster and a PDF document for sharing in virtual groups. They provided explanations regarding the program definition, objectives, student responsibilities, responsibilities of those being shadowed, and rules/regulations. Additionally, a video was prepared to orient students on patient interaction, depicting a scenario of patient interaction through role-playing.
- **Determination of learning activities:** The intended learning activities for students included performing the following tasks (the complete set of questions for each is provided provided in appendix 1, supplementary file:
- **Nurse shadowing:** Conducting an interview with a nurse and observing their activities during a 4-hour presence in the hospital.
- **Patient shadowing:** Taking a non-medical history during patient shadowing over a 4-hour presence in the hospital.
- **Intern shadowing:** Conducting an interview with an intern and observing their activities during a 2-hour presence in the hospital.
- **Health care provider shadowing:** Conducting an interview with a health care provider and observing their activities during a 2-hour presence in comprehensive health centers.
- **Development of student assessment tool:** Following the previous step, a tool for assessing students was designed, consisting of a set of questions based on reflective models, specifically Gibbs’ Reflective Cycle. This model included 6 categories of questions for each shadowing stage: 1) Description of the experience (place, time, etc.), 2) Student’s feelings during the experience, 3) Positive and negative aspects of the experience, 4) Lessons learned from the experience, 5) Suggestions for improving the experience, and 6) Incidental events during shadowing. These questions were included in their final exam.
- **Development of program evaluation tool:** A student feedback questionnaire was also designed by the executive team. This tool included demographic information and a list of shadowing objectives. Students were asked to rate the extent to which the shadowing helped them achieve each objective on a Likert scale from ‘very little’ to ‘very much’.

#### 4. Implementation

The program was implemented for all 799 medical students. The shadowing program was implemented in the first semester of the 2024-2025 academic year (starting September 2024) for all 799 basic sciences medical students. The program took place in diverse clinical environments, including teaching hospitals and comprehensive health service centers across Isfahan province. Second-semester students participated in nurse shadowing for 4 hours in teaching hospitals. Third-semester students undertook the patient shadowing experience in specialized hospital wards housing patients with COPD, heart failure, and diabetes. This selection was due to the chronic nature of these diseases and their suitability for basic sciences students’ understanding. Fourth-semester students participated in intern shadowing for 2 hours in hospital emergency departments. Fifth-semester students participated in health care provider shadowing at comprehensive health centers in Isfahan city and surrounding towns. The selection of these settings was based on criteria including alignment with the educational objectives of each semester, a history of satisfactory collaboration with the healthcare staff, and practical considerations such as distance from the city center or university.

The program was implemented in September 2024 within the framework of the “Medical Ethics” course for all basic sciences students (semesters 2-5). This course was chosen due to the alignment of its objectives with the shadowing program, as the Medical Ethics course focuses on familiarizing students with the professional path of medicine and practicing communication and professional skills.

Program implementation involved extensive coordination. Students were grouped individually or in pairs based on their availability for attendance at hospitals and health centers. Each group was assigned to shadow one individual (nurse, patient, intern, or health care provider) for 2 to 4 hours.

To ensure implementation quality, a comprehensive and multi-faceted orientation process was designed. Separate orientation sessions were held for educational supervisors and health care providers. Pamphlet guides along with official coordination letters were provided to all individuals being shadowed. A one-hour orientation session was also held for students, covering the program objectives, rules, and especially important ethical considerations. Furthermore, to prepare third-semester students for taking non-medical histories, an instructional video using role-play was produced and made available to them.

#### 5. Evaluation

To assess the program’s effectiveness, two evaluation methods were used. The primary tool for student assessment was the “Reflection Form” designed based on Gibbs’ six-stage model (13), which students completed and uploaded to the exam system as part of their end-of-semester assessment. Alongside this, a “researcher-made feedback questionnaire” was used to evaluate the overall program. This questionnaire first collected student demographic information (age, gender, semester) and then included items related to the course objectives. Students were asked to indicate the level of achievement of the program’s learning objectives on a Likert scale (from ‘Very Little’ to ‘Very Much’). This questionnaire was distributed via Google Forms through student representatives, and completion was voluntary.

Data from both tools were collected and analyzed using SPSS software (version 26). To move beyond merely descriptive reporting and better utilize the large sample size (n=799), we conducted both descriptive and inferential analyses. Descriptive statistics (frequency, mean, standard deviation) were calculated for all questionnaire items. Additionally, independent t-tests were performed to compare mean scores between gender groups and across different semesters (2–5), with statistical significance set at p<0.05. Effect sizes (Cohen’s d) were calculated for significant differences (14). We explicitly acknowledge the lack of a control group and a pre-test as major limitations (15); therefore, causal inferences cannot be drawn. The following results should be interpreted as descriptive and exploratory. This process aimed to measure the achievement of educational objectives and identify program strengths and weaknesses for improving future iterations.

### Reporting guidelines

The manuscript has been prepared following the SQUIRE 2.0 (Standards for QUality Improvement Reporting Excellence) guideline (16). The completed SQUIRE checklist is provided as a supplementary file (Appendixes 2).

### Ethical considerations and program monitoring

Given the program’s sensitivity, especially in the “Patient Shadowing” component, significant emphasis was placed on adhering to ethical principles. Students were specifically instructed on the necessity of maintaining confidentiality and respecting patient privacy; these points were also detailed in the program’s written guide. For continuous monitoring and implementation support, a communication channel was established via social media with student representatives. This mechanism allowed for the prompt identification and resolution of problems during implementation and gathered valuable feedback for future program optimization.

### Findings

The research findings based on the program implementation stages are as follows:

### Analysis Phase

In this phase, audience analysis was conducted. To analyze the audience, characteristics related to age, gender, number, semester, and completed courses/training needed to be extracted. This information was obtained by reviewing the Ham Ava educational system and through interviews with the medical faculty’s education expert (Table 1).

**Table 1:** Extracted Student Information.

| Course | Number of Students | Number of Males | Number of Females |
| --- | --- | --- | --- |
| Medical Semester 2 | 227 | 105 | 122 |
| Medical Semester 3 | 212 | 92 | 120 |
| Medical Semester 4 | 187 | 93 | 94 |
| Medical Semester 5 | 173 | 95 | 78 |

### Design phase

Based on the results of the previous phase, a precise and coherent educational program was designed for each semester. This program, which was ultimately implemented, is summarized in Table 2.

**Table 2:**
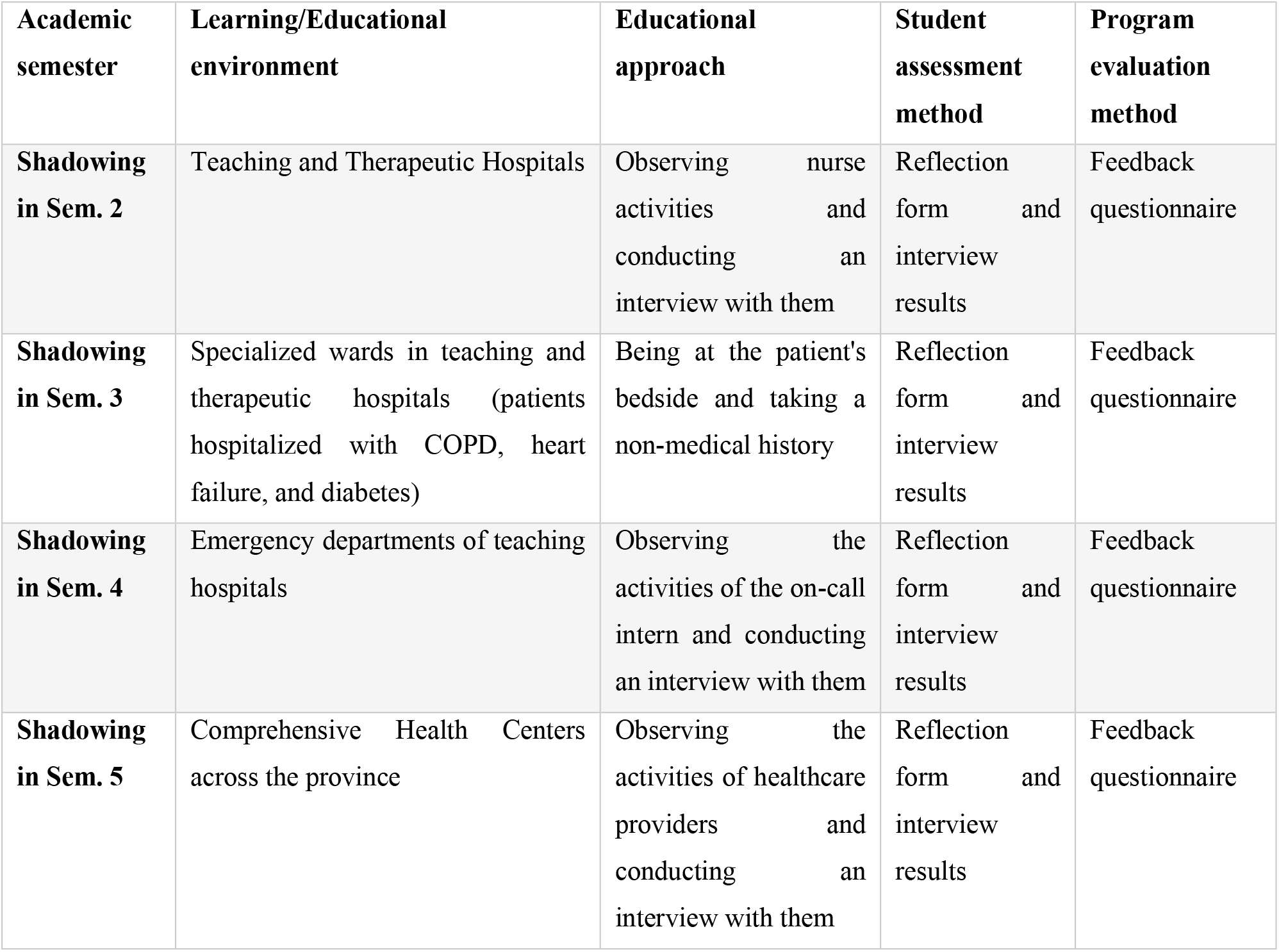
Summary of the designed shadowing program.

### Development phase

All required educational content and support tools were produced and prepared. The tangible outcomes of this phase included the preparation of four sets of educational guides for different participant groups (students, those being shadowed, supervisors, and education deputies), an instructional video using role-play, and two main assessment tools (the Gibbs model-based Reflection Form and the researcher-made feedback questionnaire).

### Implementation phase

The program was successfully operationalized with complete coverage for all 799 target students. The program was implemented in 12 teaching hospitals and comprehensive health centers across Isfahan province, with each student participating in the practical shadowing experience for 2 to 4 hours.

### Evaluation phase

Two tools were used to assess the program’s effectiveness. The feedback questionnaire was completed by 650 students (response rate: 81.4%). To move beyond purely descriptive reporting and better utilize the large sample size (n=799 for implementation, n=650 for questionnaire), we conducted both descriptive and inferential statistical analyses. All outcomes are based on student self-reported perceptions, which do not directly measure objective learning or behavioral change. Therefore, results should be interpreted as subjective evaluations of the program’s immediate impact. Given the absence of a control group and a pre-test, causal inferences cannot be drawn from these findings.

### Descriptive findings

The results from the questionnaire analysis are presented in Tables 3 to 5. In the nurse shadowing program, based on the analysis results, “Creating motivation to strive to become a good doctor” (Mean=3.95, SD=1.2), “Paying more attention to personal weaknesses and characteristics” (Mean=3.77, SD=1.2), and “Developing a positive perspective towards the clinical environment” (Mean=3.80, SD=1.3) received the highest scores, respectively. In contrast, the lowest mean was related to “Willingness to attend on holidays” (Mean=2.31, SD=0.8). Full details are provided in Table 3.

**Table 3:**
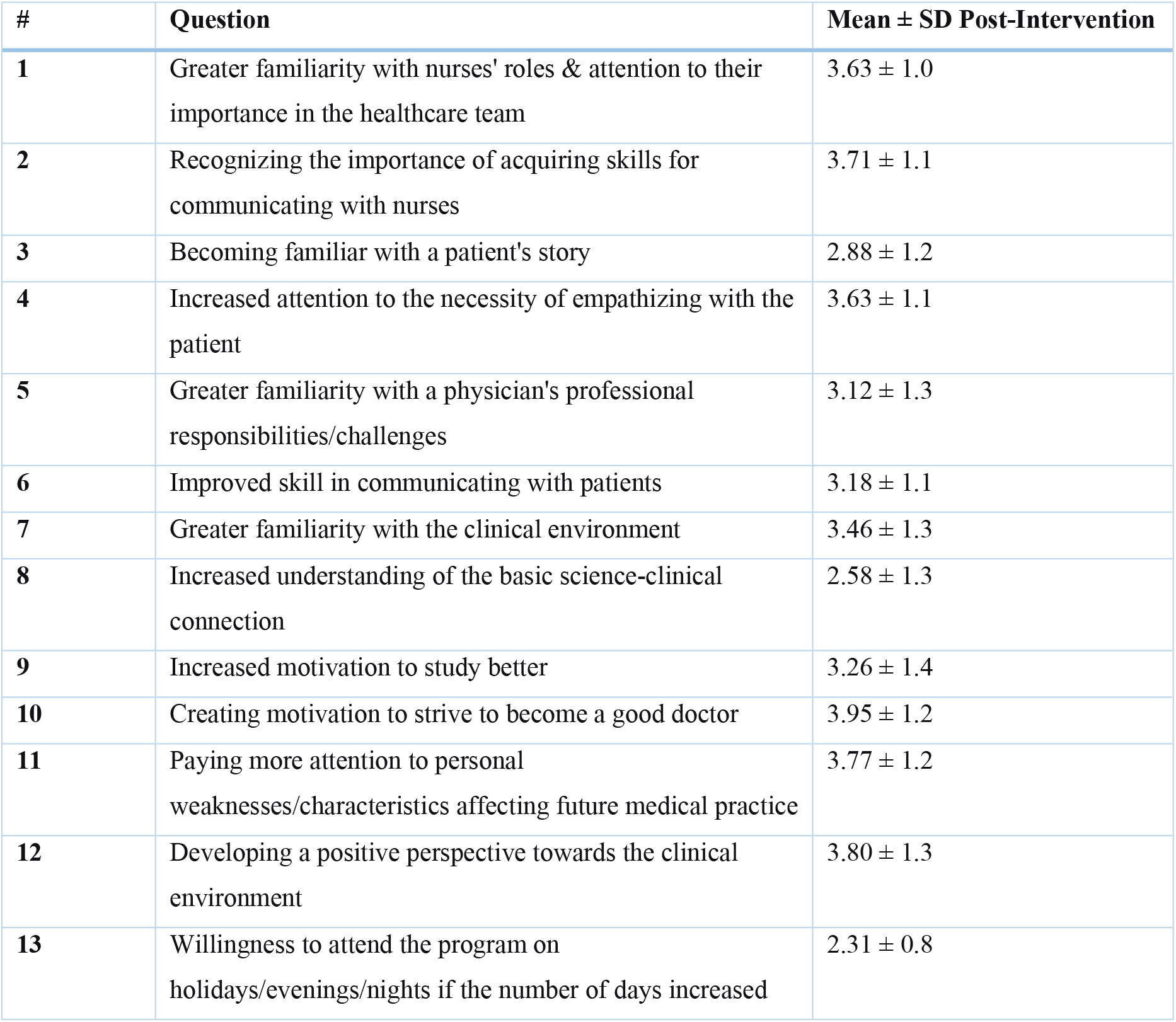
Student feedback results for the nurse shadowing program post-intervention.

| # | Question | Mean ± SD Post-Intervention |
| --- | --- | --- |
| 1 | Greater familiarity with nurses' roles & attention to their importance in the healthcare team | 3.63 ± 1.0 |
| 2 | Recognizing the importance of acquiring skills for communicating with nurses | 3.71 ± 1.1 |
| 3 | Becoming familiar with a patient's story | 2.88 ± 1.2 |
| 4 | Increased attention to the necessity of empathizing with the patient | 3.63 ± 1.1 |
| 5 | Greater familiarity with a physician's professional responsibilities/challenges | 3.12 ± 1.3 |
| 6 | Improved skill in communicating with patients | 3.18 ± 1.1 |
| 7 | Greater familiarity with the clinical environment | 3.46 ± 1.3 |
| 8 | Increased understanding of the basic science-clinical connection | 2.58 ± 1.3 |
| 9 | Increased motivation to study better | 3.26 ± 1.4 |
| 10 | Creating motivation to strive to become a good doctor | 3.95 ± 1.2 |
| 11 | Paying more attention to personal weaknesses/characteristics affecting future medical practice | 3.77 ± 1.2 |
| 12 | Developing a positive perspective towards the clinical environment | 3.80 ± 1.3 |
| 13 | Willingness to attend the program on holidays/evenings/nights if the number of days increased | 2.31 ± 0.8 |

In the physician shadowing program, data analysis showed that “Creating motivation to strive to become a good doctor” (Mean=3.75, SD=1.2), “Improved skill in communicating with patients” (Mean=3.53, SD=1.2), and “Increased attention to the necessity of empathizing with the patient” (Mean=3.50, SD=1.2) were the three items with the highest means. The lowest score was again for “Willingness to attend on holidays” (Mean=2.09, SD=0.9). Complete data is in Table 4.

**Table 4:** Student feedback results for the physician shadowing program post-intervention.

| # | Question | Mean ± SD Post-Intervention |
| --- | --- | --- |
| 1 | Greater familiarity with nurses' roles & attention to their importance in the healthcare team | 3.60 ± 1.1 |
| 2 | Recognizing the importance of acquiring skills for communicating with nurses | 3.47 ± 1.2 |
| 3 | Becoming familiar with a patient's story | 3.73 ± 1.0 |
| 4 | Increased attention to the necessity of empathizing with the patient | **3.50 ± 1.2** |
| 5 | Greater familiarity with a physician's professional responsibilities/challenges | 3.45 ± 1.2 |
| 6 | Improved skill in communicating with patients | **3.53 ± 1.2** |
| 7 | Greater familiarity with the clinical environment | 3.48 ± 1.3 |
| 8 | Increased understanding of the basic science-clinical connection | 2.53 ± 1.3 |
| 9 | Increased motivation to study better | 2.81 ± 1.4 |
| 10 | Creating motivation to strive to become a good doctor | **3.75 ± 1.2** |
| 11 | Paying more attention to personal weaknesses/characteristics affecting future medical practice | 3.32 ± 1.2 |
| 12 | Developing a positive perspective towards the clinical environment | 3.18 ± 1.3 |
| 13 | Willingness to attend the program on holidays/evenings/nights if the number of days increased | 1.87 ± 0.8 |

Results for **healthcare provider shadowing** also indicated that “Creating motivation to strive to become a good doctor” (Mean=3.75, SD=1.2), “Improved skill in communicating with patients” (Mean=3.53, SD=1.2), and “Increased attention to the necessity of empathizing with the patient” (Mean=3.50, SD=1.2) were the three items with the highest means, highlighting the significant impact on student motivation and communication skills. The lowest score was again for “Willingness to attend on holidays” (Mean=2.09, SD=0.9). Table 5 details these results.

**Table 5:** Student feedback results for the healthcare provider shadowing program post-intervention.

| # | Question | Mean ± SD Intervention |
| --- | --- | --- |
| 1 | Becoming familiar with a patient's story | **3.39 ± 1.3** |
| 2 | Increased attention to the necessity of empathizing with the patient | **3.50 ± 1.2** |
| 3 | Greater familiarity with a physician's professional responsibilities/challenges | **3.47 ± 1.2** |
| 4 | Improved skill in communicating with patients | **3.53 ± 1.2** |
| 5 | Greater familiarity with the clinical environment | **3.31 ± 1.3** |
| 6 | Increased understanding of the basic science-clinical connection | **2.89 ± 1.3** |
| 7 | Increased motivation to study better | **3.11 ± 1.4** |
| 8 | Creating motivation to strive to become a good doctor | **3.75 ± 1.2** |
| 9 | Paying more attention to personal weaknesses/characteristics affecting future medical practice | **3.33 ± 1.2** |
| 10 | Developing a negative perspective towards the clinical environment | **3.14 ± 1.3** |
| 11 | Willingness to attend the program on holidays/evenings/nights if the number of days increased | **2.09 ± 0.9** |

Inferential findings: To utilize the large sample size (n=650 for feedback questionnaire), we performed independent t-tests to compare mean overall satisfaction scores (averaged across all 13 items) between different shadowing types and between gender groups.

- Comparison across shadowing types: Mean overall satisfaction was highest for nurse shadowing (Mean=3.32, SD=0.42), followed by physician shadowing (Mean=3.19, SD=0.45) and healthcare provider shadowing (Mean=3.17, SD=0.44). One-way ANOVA revealed a statistically significant difference between the three groups (F(2, 647)=4.86, p=0.008, partial η^2^=0.015). Post-hoc Tukey tests showed that nurse shadowing had significantly higher scores than physician shadowing (p=0.02) and healthcare provider shadowing (p=0.01), with small effect sizes (Cohen’s d=0.30 and 0.34 respectively).
- Comparison across genders: Female students (n=342) reported slightly higher overall satisfaction (Mean=3.29, SD=0.43) than male students (n=308, Mean=3.20, SD=0.44). An independent t-test showed a borderline significant difference (t(648)=2.21, p=0.027, Cohen’s d=0.17), indicating a very small effect.
- Item-level comparison: The item “Increased understanding of the basic science-clinical connection” consistently received low scores across all programs (mean range 2.53– 2.89). An independent t-test comparing this item between nurse shadowing (Mean=2.58) and physician shadowing (Mean=2.53) showed no significant difference (p=0.48).

Given that all data are perception-based and no objective pre-post or control group exists, causal conclusions cannot be drawn. These inferential results should be considered exploratory.

### Overview of findings

An overview of all three tables identifies “Creating motivation to become a good doctor” in two programs (Nurse and Healthcare Provider shadowing) and “Empathy with the patient” in the Physician shadowing program as the strongest points of the program. Conversely, “Increased understanding of the basic science-clinical connection” received relatively low scores across all programs. The reported standard deviations (mostly between 0.8 and 1.4) indicate a relatively balanced dispersion of opinions around the mean.

## Discussion

This study demonstrates that a shadowing program systematically designed using the ADDIE model enhanced multiple dimensions of medical student learning and professional attitudes. However, beyond simply confirming previous findings, this discussion aims to explain why and how the program worked from a theoretical standpoint. Drawing on Lave and Wenger’s situated learning theory (legitimate peripheral participation), we propose that the shadowing program facilitated learning through three interconnected mechanisms: (a) goal-directed observation of clinical role models, (b) structured reflection using Gibbs’ cycle (13), and (c) mini-clinical dialogues during pre-briefing and debriefing sessions. These mechanisms collectively transformed passive observation into active learning—a distinction that prior descriptive studies have largely overlooked (7 ,5).

In nurse shadowing, the strongest impact was observed on “Creating motivation to become a good doctor” and “Paying more attention to personal weaknesses and characteristics.” From a theoretical perspective, observing nurses—who are often undervalued in traditional medical hierarchies— may disrupt students’ pre-existing stereotypes about professional roles, thereby prompting critical self-reflection. This mechanism goes beyond simple role modeling and engages what Jarvis (17) termed “disjuncture” as a trigger for transformative learning. This finding aligns with Hood et al. (18), who noted that understanding diverse team roles enhances students’ motivation and systems-thinking. Furthermore, the focus on self-awareness supports Annadurai et al. (19), who identified reflection as a cornerstone of competency-based education.

Conversely, physician shadowing most prominently enhanced empathy and understanding of the patient’s story. The direct exposure to patients’ narratives appears to activate narrative competence—the ability to acknowledge, absorb, interpret, and act on stories of illness (20). This mechanism is qualitatively different from simple observational learning because it requires emotional engagement and perspective-taking. This is particularly crucial given the documented decline in empathy during medical training, as quantified by Shakurnia et al. (21). The value of this narrative-based approach is further supported by Lam et al. (22), who found that life story interviews help students connect with patients. However, as Milota et al. (23)caution, more robust evidence is needed to confirm its impact on long-term behavioral change. Our study contributes to this gap by providing mechanism-based rather than merely correlational evidence.

In healthcare provider shadowing, the highest scores were for “Increased motivation to become a good doctor” and “Improved patient communication skill.” We theorize that observing primary care providers—who manage continuity, prevention, and chronic illness—exposes students to the “longitudinality” of care, a dimension absents in hospital-based shadowing. This exposure may strengthen students’ understanding of patient-centeredness as an ongoing relationship rather than a single encounter. This aligns with Henschen et al. (24), who associated longitudinal clinical exposures with improved patient-centeredness and continuity of care.

A notable cross-cutting finding was the consistently low score for “Increased understanding of the basic science–clinical connection.” This finding is theoretically important: it suggests that shadowing, in its current design, promotes affective and social learning (motivation, empathy, self-awareness) but not cognitive integration. According to Kulasegaram et al. (3) , basic science knowledge must be activated in clinical contexts through deliberate cuing—a mechanism absents in passive observation. This gap is well-documented; Kulasegaram et al. (3) noted a lack of rigorous evaluation for integration strategies, and Kercheval et al. (2) highlighted student struggles in perceiving this relevance, especially early in training. Our study adds to this literature by empirically demonstrating that shadowing alone is insufficient for cognitive integration, thereby identifying a specific design flaw to be addressed in future iterations (e.g., adding pre-shadowing basic science review sessions).

When compared, the three shadowing experiences reveal a complementary educational pattern. Nurse shadowing fostered self-awareness, physician shadowing cultivated empathy, and primary care shadowing enhanced systems understanding and motivation. Rather than merely describing this pattern, we argue that it reflects a multidimensional professional identity formation process, where different role models activate different facets of the “possible self” (25). This triangulated approach is supported by Yang et al. (5), who found that varied shadowing experiences help students perceive diverse competencies. Furthermore, Hilton & Jerjes (8) demonstrated that structured, multi-phase shadowing can significantly improve key attributes like empathy and patient-centeredness. Critically, these experiences are not interchangeable; together, they create a multidimensional educational paradigm that comprehensively supports professional growth. This study’s novel contribution is not merely confirming that shadowing works, but specifying which type of shadowing works for which outcome—a level of granularity absent in prior descriptive studies(8)(7) .

Finally, the uniformly low willingness to attend on holidays points to a need for future programs to better accommodate student work-life balance. From an implementation science perspective, this finding highlights the importance of considering student burnout risk even in short-term educational interventions (11).

### Strengths and limitations

This study possesses several notable strengths. The systematic application of the ADDIE instructional design model provided a structured and evidence-based framework for the program’s development, implementation, and evaluation. The intervention demonstrated high scalability and feasibility, successfully engaging all 799 target students across 12 diverse clinical settings. The use of multiple assessment methods, including Gibbs-based reflection forms and a validated questionnaire, ensured robust and triangulated data collection. Furthermore, the program’s progressive design, which exposed students to various healthcare roles, fostered holistic professional development, while a strong emphasis on ethics and comprehensive orientation ensured its integrity and safety.

However, several limitations should be acknowledged. The study’s conduct within a single university may restrict the generalizability of its findings. The primary reliance on self-reported data introduces a potential for social desirability bias. More importantly, the absence of a control group and a pre-test design means that causal attributions cannot be made—a limitation explicitly noted in the Methods section. The relatively low scores concerning the integration of basic and clinical sciences indicate a curricular gap that needs addressing. Challenges with student scheduling flexibility were evident. Future studies should employ quasi-experimental designs with control groups and objective pre-post measures of learning (e.g., knowledge tests or observed clinical skills) to strengthen causal inference.

### Suggestions for future research

This study opens several promising avenues for further investigation. Future research should explore the long-term impact of this shadowing intervention by tracking participants into their clinical years and residency, examining its sustained effects on professional identity formation, clinical competencies, and career choices. Additionally, a comprehensive qualitative analysis of student reflection forms would provide deeper insights into the transformative aspects of their learning experiences and the development of specific professional attributes. Further investigation into the dynamics of student interactions with various healthcare team members, particularly nurses and patients, could illuminate how these relationships influence professional socialization and interprofessional collaboration skills. These research directions would substantially enhance our understanding of how early clinical exposures shape medical trainees’ professional development.

## Conclusion

Based on the findings of this study, the shadowing program designed using the ADDIE model demonstrated its effectiveness in developing various professional dimensions of medical students. The results indicate that each type of shadowing complementarily impacted different aspects of learning; nurse shadowing had the greatest impact on self-awareness and professional motivation, physician shadowing on strengthening empathy and patient understanding, and healthcare provider shadowing on understanding the health system and communication skills. However, the relative weakness in linking basic sciences with clinical practice indicates that this program needs intentional reinforcement in establishing this connection. Overall, the findings confirm that incorporating structured shadowing programs into the curriculum can significantly contribute to nurturing well-rounded physicians with essential soft skills.

## Abbreviations

ADDIE: Analysis Design Development Implementation Evaluation

## Clinical trial number

Not applicable.

## Supplementary information

The student’s questions regarding the determination of learning activities are provided in Appendix 1, and the SQUIRE 2.0 checklist is provided in Appendix 2.

## Authors’ contributions

O.A. and CH.T. contributed to the conceptualization, methodology, project administration, supervision, and validation of the study. S.GH. and O.A. and Z. Kh. and K. H. and A.A and MH. F. and M.Y. performed data curation, formal analysis, investigation, and resources collection, and wrote the original draft. CH.T and O. A. contributed to writing through review and editing. All authors have reviewed and approved the final manuscript.

## Data availability statement

Data is provided within the manuscript.

## Declarations

### Ethics approval and consent to participate

This study was approved by National Agency for Strategic Research in Medical Education (Ethics Code: IR.NASRME.REC.1402.086). All research procedures followed the university’s ethical regulations and the Declaration of Helsinki. We obtained informed consent from every participant, protected the confidentiality of their information, and upheld their right to withdraw from the study without penalty.

### Consent for publication

Not applicable.

